# The RESIST Senior Individuals Cohort: Design, participant characteristics and aims

**DOI:** 10.1101/2024.04.29.24306533

**Authors:** LM Roesner, KM Gupta, V Kopfnagel, N van Unen, Y Kemmling, J Heise, S Castell, X Jiang, L Riemann, S Traidl, B Lange, KW Sühs, T Illig, T Strowig, Y Li, R Förster, J Huehn, TF Schulz, T Werfel, the RESIST SI Cohort Investigators

**Author notes:** equal contribution (LM Roesner, KM Gupta). equal contribution (TF Schulz, T Werfel).

## Abstract

The number of older adults worldwide is growing exponentially. However, while living longer, older individuals are more susceptible to both non-infectious and infectious diseases, at least in part due to alterations of the immune system. Here, we report on a prospective cohort study investigating the influence of age on immune responses and susceptibility to infection. The RESIST Senior Individuals (SI) cohort was established as a general population cohort with a focus on the elderly, enrolling an age- and sex-stratified sample of 650 individuals (n=100 20-39y, n=550 61-94y, 2019-2023, Hannover, Germany). It includes clinical, demographic, and lifestyle data and also extensive biomaterial sampling. Initial insights indicate that the SI cohort exhibits characteristics of the aging immune system and the associated susceptibility to infection, thereby providing a suitable platform for the decoding of age-related alterations of the immune system and unraveling the molecular mechanisms underlying the impaired immune responsiveness in aging populations by exploring comprehensive, unbiased multi-omics datasets.

## Introduction

Due to steady improvements in the healthcare system, human life expectancy is increasing^1^. At present, ∼500 million people are above the age of 65 globally and this number is expected to increase to ∼1.5 billion by 2050^2^. However, enhancement of lifespan does not guarantee enhancement in healthspan, i.e., the life duration free from chronic diseases and disabilities^3^. Elderly individuals are more prone to both non-infectious and infectious diseases mainly because of the age-associated physiological dysfunctions^4,5^ and immune system remodeling^3^.

The particular risk for fatal infection of the respiratory tract caused by pathogens in elderly individuals has been observed during the SARS-CoV-2 pandemic: elderly individuals were found to be highly susceptible to develop severe forms of COVID-19 with very high case fatality rates^6^. Further, latent virus infections such as Varicella zoster virus (VZV) re-activate more often with increased age, and can lead to severe disease, including acute and/or long-term complications^7^. Being considered as the orchestrating system of the body to maintain health, the immune system has the task of fighting pathogens. However, if it is not properly adapted, this can lead to diseases in especially the aging organism. The inability to mount appropriate immune responses seems to be a central contributing factor to increased mortality rates in the elderly population. However, the immune system is not defective per se, since strong immune responses to e.g. VZV vaccines are observed even in subjects older than 80 years of age^8^.

Aging of the immune system is a highly complex process, and several factors contribute to impaired immunity in elderly individuals. As a consequence of the thymic involution and parallel acute and chronic antigenic stress over the lifetime^9,10^, an absolute decrease of naive T cell numbers is observed, leading to a reduced diversity of the TCR repertoire. Nevertheless, the thymus is capable of producing naive T cells also in high age, and thymus-independent mechanisms ensure a functional adaptive immune system in homeostasis. The adaptive immune system also undergoes constant changes in parallel, e.g. via homeostatic proliferation of naïve T cells or stem cell-like memory T cells induced by cytokines^11^. On the other hand, frequencies of memory T cells, primed and stimulated to undergo proliferation by their cognate antigens, are relatively increased. A prominent example are T cells specific for human cytomegalovirus (CMV), which are drastically increased in infected, aged individuals^12,13^, presumably due to the recurring re-activations of this latent infection. Furthermore, T cells of elderly individuals have been reported to be energy deprived due to metabolic differences^14^ and vaccination studies in elderlies reported reduced T cell reactivity and expression of cellular markers for exhaustion^15^. The aged innate immune system shows a general tendency to produce higher levels of inflammatory mediators, including prostaglandin E2, interleukin(IL)-6, interferon(IFN)-γ and TNF-α. This phenomenon is often termed inflammaging^16^. It is under debate whether this may lead to multimorbidity, cardiovascular diseases, and frailty^17^.

Studies on longevity show that not only the triggering of the immune system by pathogens, but also lifestyle, nutrition and the microbiome are important factors that shape the immune aging over the years^18,19^. Adding further complexity is the fact that each person’s immunological response to a given stimulus or pathogens is unique. Thus, different people with the same infectious disease may present with varying degrees of severity, vaccines can elicit different immune responses in individuals, and different people may show varying responsiveness to the same medical therapy^20^. This in turn indicates that aging of the immune system is an individual process^21^ which requires novel concepts of personalized medicine in the prevention and management of infectious and immunological-driven diseases in elderly individuals. To consider as many factors as possible, multi-omics investigations are needed to characterize the complex systems of the immune network and also the inter-individual variance of immune responses.

### Aims

Considering the above, the RESIST senior individuals (SI) cohort was set up as a general population cohort to (i) describe and investigate age-related differences and changes in the human immune system, in particular innate and adaptive immune responses to pathogens, (ii) investigate the influence of environmental, lifestyle and dietary factors on the shaping of the immune system in old age, (iii) explore the role of the intestinal microbiome in shaping the immune system in old age and (iv) compare immune responses and the susceptibility to virus infection/reactivation of aged healthy individuals with those of patients suffering from recurrent VZV and herpes simplex virus (HSV) reactivations (the RESIST VZV cohort and HSV/AD cohort, see below). In brief, we aim to decode age-related alterations of immune responsiveness and identify the key players of effective immunity within aging populations by exploring comprehensive unbiased genome-wide sequencing, immune phenotyping, and multi-omics approaches (Figure 1).

**Figure 1:**
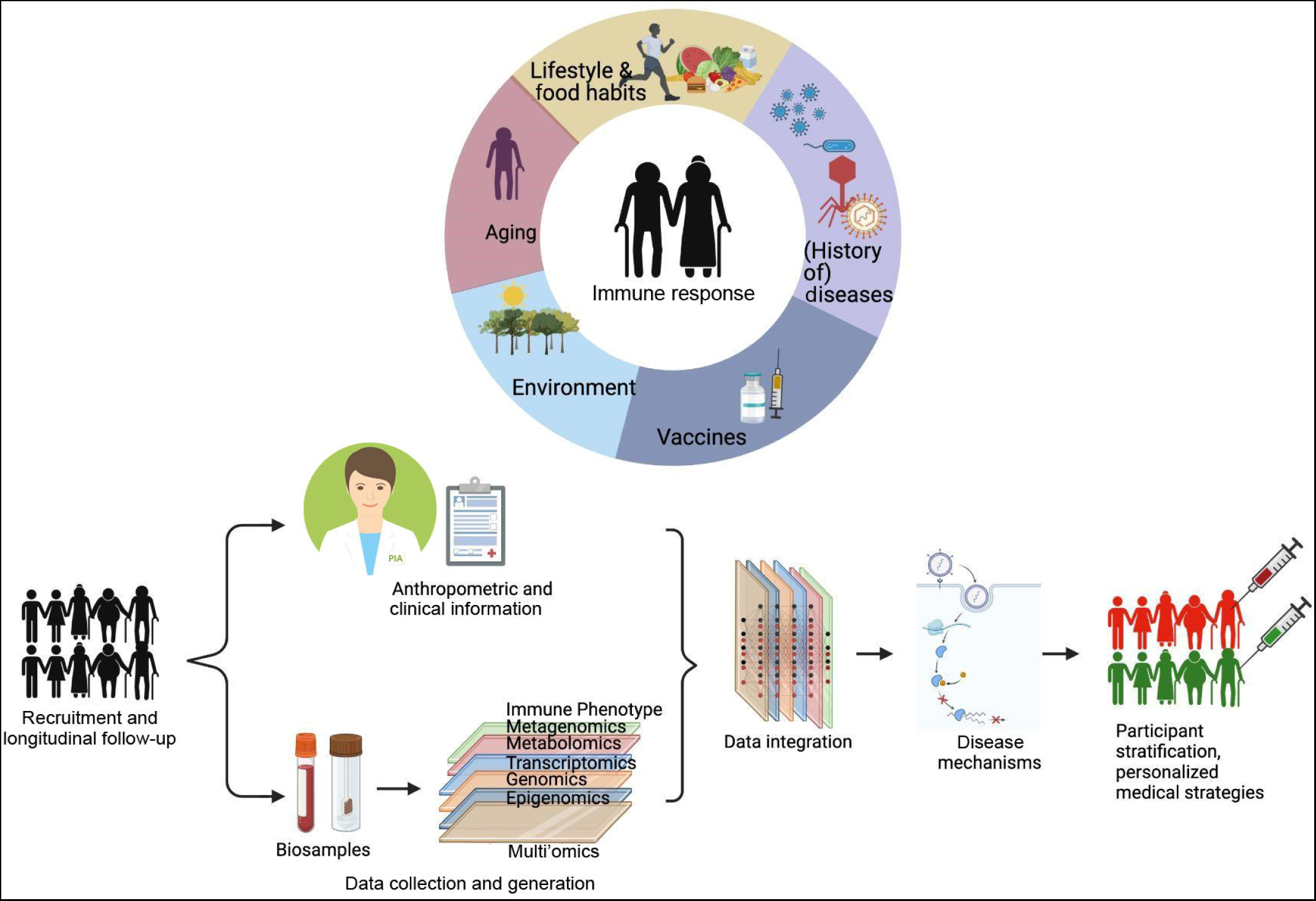
Recruitment, sampling and analysis scheme of the SI cohort.

## Results

### Study design, cohort recruitment and description of the study visits program

We conducted a prospective cohort study investigating the influence of age on the immune response to pathogens and the etiology of increased susceptibility to viral infections in old age.

The RESIST consortium (Cluster of Excellence EXC 2155), funded within the framework of the “Federal and State Excellence Strategy” by the German Research Foundation (DFG), investigates the impact of genetic and life style factors as well as age on infection susceptibility. The study was approved by the local ethics committee of Hannover Medical School (MHH, see Online Methods) and participants gave their written informed consent.

The recruitment lasted from December 2019 until March 2023 and the follow-up phase was completed at the end of 2023. We recruited 650 cohort participants, stratified by age and gender, randomly selected from the residents’ registration office in the study region of Hannover, Germany. Men and women were included in equal numbers. The final overall response to written invitations from the study center was 8.4%. During the recruitment process, participants were invited and first underwent a telephone interview. Individuals were excluded from the study if they were unable to provide informed consent, and/or were not being able to understand the study information, unable to respond to interview or study context questions, and/or were unable to participate in the majority of the planned examinations. Further exclusion criteria were reception of any organ transplant, current pregnancy, or current anti-inflammatory or immunosuppressive systemic medications such as glucocorticosteroids, cytostatic drugs, cyclophosphamide, biologics / antibodies targeting inflammatory mediators, mTOR inhibitors, JAK inhibitors, or azathioprine.

Suitable participants were invited to the study center to participate in a standardized personal face-to-face interview, to complete self-administered questionnaires and to undergo several standardized physical and medical examinations all of which were documented in the eResearch System “Prospective Monitoring and Management – App” (PIA)^22^. In addition, all participants provided blood, 561 also provided stool samples, and 109 contributed nasal swabs. All participants received a letter listing a selected set of laboratory data in the weeks following their study visit. There was no financial compensation for participation apart from reimbursement of travel costs or taxi voucher and a free lunch.

650 participants (Figure 2) with a focus on the age group of 60-100 years (“Elderly”, n=550) were included from December 2019 - March 2022. A subgroup of n=100 20-39 year-olds (“Young adults”) was included to explore age-related differences.

**Figure 2:**
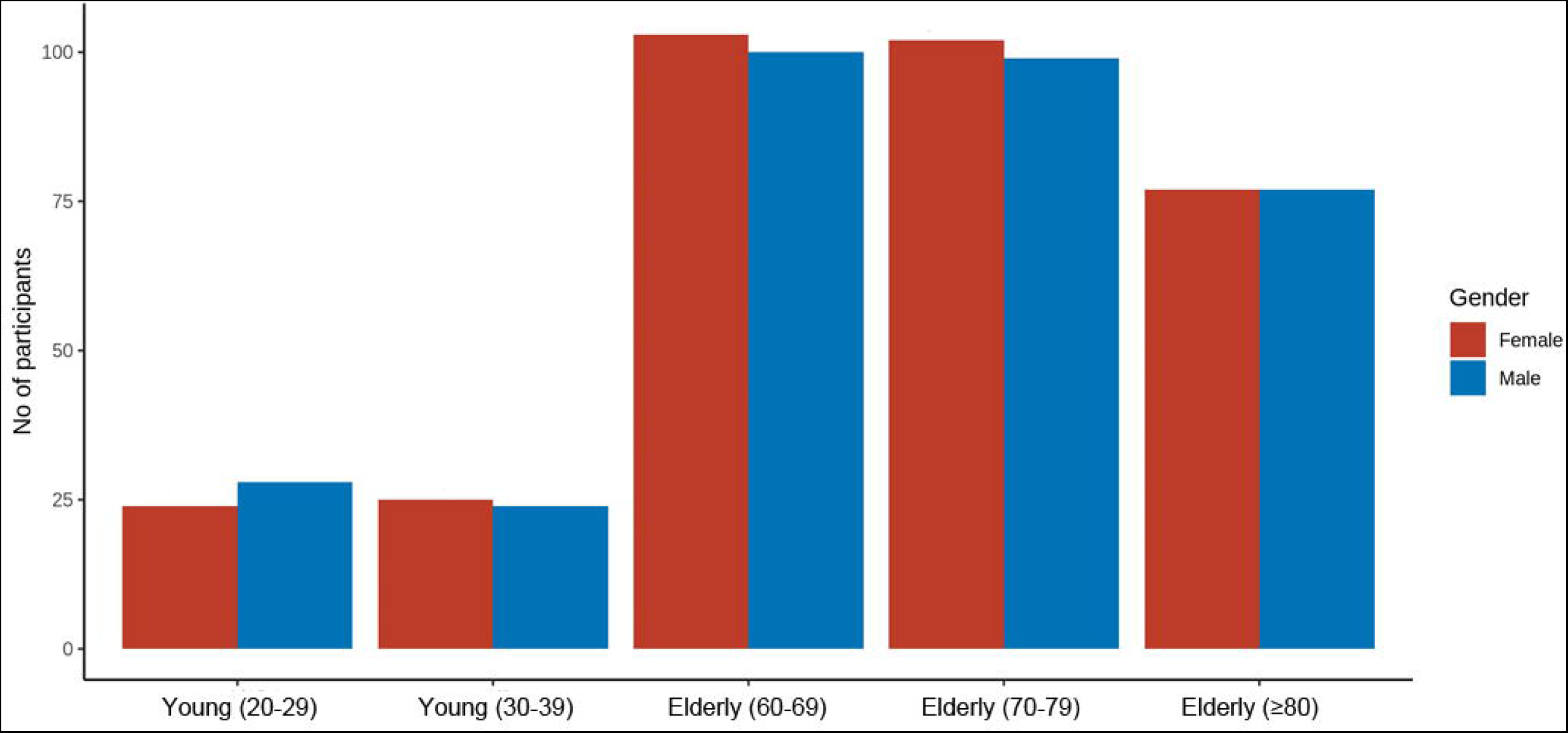
Age and gender profile of participants. Age subgroups of “Young adults” and “Elderly” are depicted in brackets.

250 of the elderly participants participated in a second visit to the study center 1.5-2.5 years after their first study visit, which included the same examinations and biospecimens as performed during the first study visit.

All SI cohort participants followed a 2.5 h recruitment protocol including questionnaire assessments and basic physical and medical examinations. To allow comparisons with the largest epidemiological cohort study so far ever conducted in Germany, the German National Cohort (GNC, ‘NAKO Gesundheitsstudie’), we implemented the following instruments with permission of GNC^23^: Interview (modified), hand grip strength anthropometry, blood pressure and heart rate, medication use, COVID-19 questionnaire. This also facilitated the use of instruments relevant to cohort research in Germany that have already been demonstrated to be applicable. The Cluster of Excellence RESIST signed an agreement for the utilization of instruments from the GNC for the SI cohort. The partners agreed to collaborate regarding the utilization of instruments, questionnaires and standard operating procedures (SOPs) of GNC, created and developed for the GNC.

The standardized personal face-to-face 271-item interview included questions on general life circumstances, education, lifestyle factors such as smoking and dietary behaviors, and clinical information, e.g. history of cardiovascular diseases, malignancies, metabolic disorders, musculoskeletal diseases, lung diseases, allergies, gastrointestinal or liver diseases, skin diseases, kidney diseases, neurological and psychiatric disorders, eye diseases, undergone surgeries, and – with a particular focus - infectious diseases

The self-administered standardized questionnaires contained Beck Depression Inventory (BDI)-Fast Screen, Barthel index (BI, only participants ≥ 60 years), a fitness questionnaire (only participants <40 years), and nutrition questionnaires. The interview was followed by an assessment of vaccinations, current medication, and allergies. Throughout the clinical investigations basic anthropometric data, e.g. height (cm), waist (cm), & weight (kg), blood pressure (both diastole & systole), heart rate, BIA (bioelectrical impedance analysis), a skin screen, assessment of grip force, ‘Timed Up and Go’ (TUG, only participants ≥ 60 years), and Montreal Cognitive Assessment (MoCA) (only participants ≥ 60 years) was collected by an intensively trained and certified study team (Table 1).

**Table 1:** Baseline characteristics. Two-tailed χ² test of independence (smoking behaviour, migratory background) or Wilcoxon test (body mass index (BMI), height, weight, waist circumfence) were performed where applicable, comparing the groups of young adults and elderlies. n.s. = no significant difference.

| | Total | Young adults (20-39 years at baseline) | Elderly ($\geq 60$ years at baseline) | p-value |
| --- | --- | --- | --- | --- |
| Number of participants | 650 | 100 | 550 |  |
| Age (years) median (range) | 70 (22-94) | 29 (22-38) | 73 (61-94) |  |
| Gender (Female/Male) | 325 / 325 | 48/52 | 277/273 |  |
| BMI (kg/m<sup>2</sup>), median (IQR) | 25 | 24 (21-25) | 26 (23-29) | < 0.001 |
| Current smokers | 70 (10.8%) | 15 (16%) | 55 (10%) | n.s. |
| Migratory background | 10 (1.6%) | 3 (3%) | 7 (1.3%) | n.s. |
| Height (cm), median (range) | 169.4 (132.5-199.5) | 173.4 (153.9- 198.1) | 169.1 (132.5- 199.5) | < 0.001 |
| Weight (kg), median (range) | 74.5 (42.1-131.1) | 68.45 (46.8- 107.9) | 75.30 (42.1-131.1) | 0.026 |
| Waist circumference (cm), median (range) | 91.3 (62.0-144.5) | 77.85 (63.1- 113.2) | 94.20 (62.0- 144.5) | < 0.001 |

Within the framework of an additional project “App-based Infection Assessment in RESIST (iAR)”, all participants of the SI cohort who visited the study center for the first study visit, were asked to additionally participate in an app-based monitoring of infectious diseases using PIA. In this intensified infection module, which was designed to be directly comparable to the ZIFCO study^24^, a subgroup of 218 participants volunteered to participate. They received weekly questionnaires to allow reporting on acute infections of the previous week. In addition, participants were able to report symptoms immediately through a “spontaneous report”. In case of respiratory symptoms, the participants sent nasal swabs for viral diagnostics to the Institute of Virology of MHH. These participants were tracked from March 2021 through the end of 2023.

### Collection and storage of bio-specimens

To ensure comprehensive, consistently high-quality biospecimens throughout the project, biospecimen collection was carried out in cooperation with the Hannover Unified Biobank (HUB). Biospecimens were processed in the HUB laboratory or the Department of Dermatology and Allergy at MHH according to HUB standards^25^. The entire process from collection of biospecimens to processing and storage is documented in the HUB’s biobank management system. The laboratory investigations included immunophenotyping of the cohort on the humoral and cellular level, as well as generation of genetic, transcriptomic, epigenomic, immunologic, proteomic, metabolomic, and stool metagenomic data sets.

Subsequently, these data are integrated and analyzed using multi-omics analyses to investigate age-related changes in the human immune system, in particular the innate and adaptive immune responses to four different TLR agonists (LPS, Pam3Cys, poly I:C, CpG) and antigens/peptides derived from VZV, HSV, CMV, Influenza, and severe acute respiratory syndrome coronavirus 2 (SARS-CoV2), respectively.

Furthermore, the influence of environmental, lifestyle and dietary factors on the shaping of the immune system in old age is assessed. Considering the impact of the recent SARS-CoV2 pandemic on human health, especially on elderly people, from August 2020 on, the SI cohort program was extended to investigate the immune response to SARS-CoV2. It is noteworthy that most participants do not have a migratory background and several data sets of the SI cohort can be directly compared with corresponding data. Some of those comparable data sets were already generated under similar conditions from younger individuals (25 – 40 years) within the “500FG cohort”^26–28^ (Radboud University Medical Center, Nijmegen, The Netherlands). Two infection disease cohorts were established in parallel with overlapping protocols and examinations (see “Connected disease cohorts”). As mentioned above, the questionnaires and medical examinations allow a direct comparison to the GNC.

### Baseline characteristics

The MoCA, a screening tool for cognitive deficits, includes memory, attention, and orientation and is therefore often used to describe cohorts of older people or those with disease. While a maximum of 30 points can be reached, the total MoCA score of the elderly individuals of the SI cohort was 25.7 ± 3.3 (mean ± SD), while 37.9% scored below the cut-off of 26/30 points. Earlier, a mean of 26.1 ± 2.5 and a rate of 31.1% below the cut-off was reported for a German-speaking group of 283 individuals of 65 years or older^29^. TUG-measurement resulted in a mean time of 9.4 (± 4.9) seconds (± SD) in the elderly individuals of the SI cohort, which compared to a meta-analysis reporting 9.4 seconds in mean for 60-99 year-olds^30^.

The questionnaire items on infectious diseases were evaluated in detail with regard to the frequency and type of infection. 38% of the participants reported a history of infectious diseases as depicted in Table 2, and as expected, this frequency was markedly higher in elderly (40.5%) versus young adults (24%). Compared to the young, the elderly reported significantly more often a history of Influenza, Herpes zoster, Hepatitis B or C, and Tuberculosis (Table 2). While the longer lifespan may explain this effect for several of these diseases, herpes zoster appeared as a disease of the advanced age also in our cohort, with a mean age of 55 years. The GNC, which was assessed based on the same interview questions but consisted at the time point of analysis of >100,000 adults with a median age of 53 and only <3% of individuals above 70 years of age, reported lower infection frequencies compared to the ≥60 year-olds of the SI cohort^31^. Precisely, hepatitis (combined frequencies for hepatitis B and C, 2.2% GNC vs. 3.3% elderly SI), herpes zoster (8.6% GNC vs. 18.2% elderly SI) and tuberculosis (1.4% GNC vs. 3.1% elderly SI) were less frequently reported. 4% of the elderly SI cohort participants reported a history of sepsis, which was slightly higher compared to the GNC.

**Table 2:**
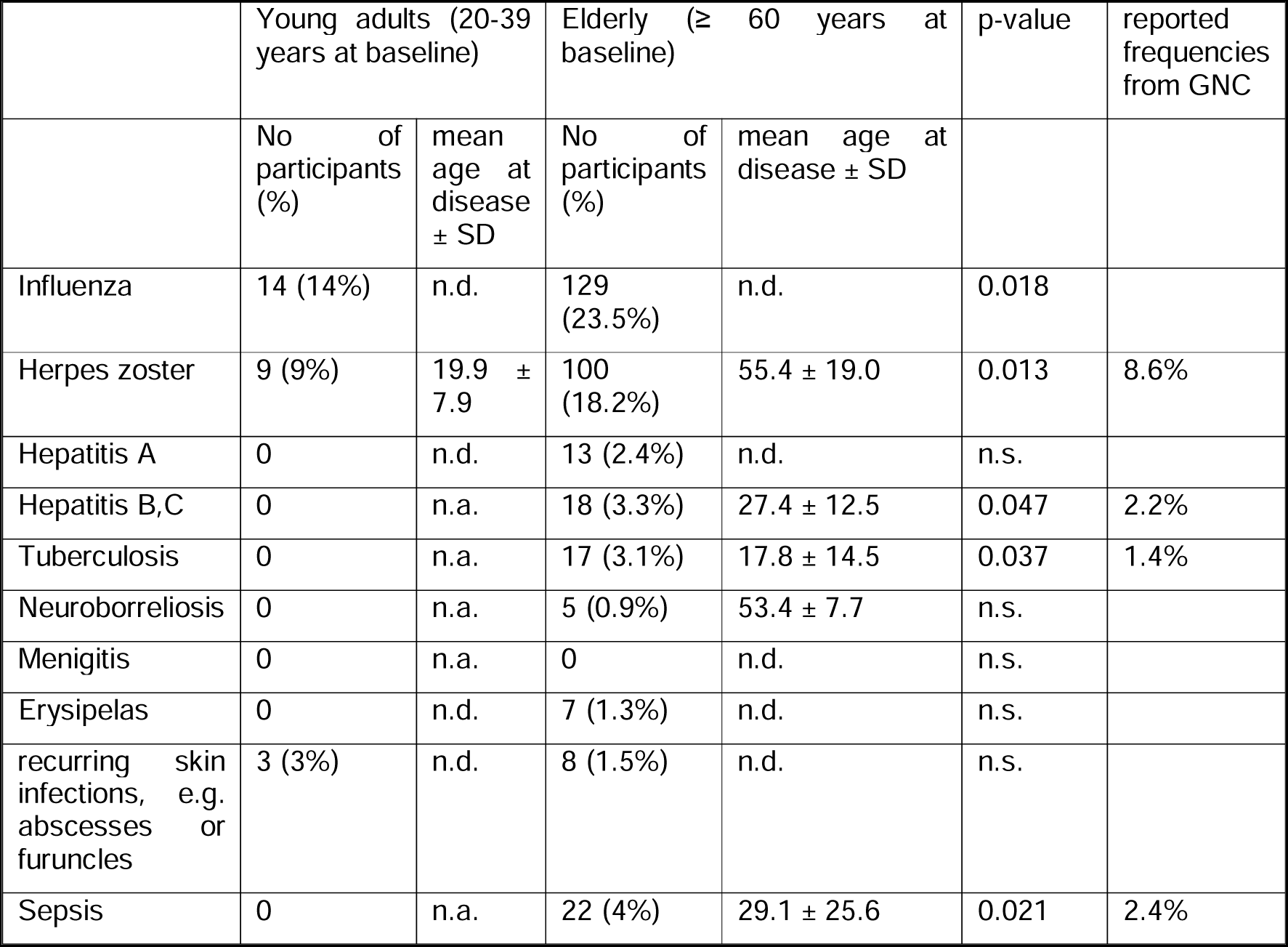
Number of subjects reporting a history of selected infectious diseases as well as sepsis. Where applicable, the mean age at which the patient suffered the disease is indicated. The hypothesis of higher infection rates in the elderly group compared to the younger was tested by one-tailed χ² test. Reported frequencies from GNC from^31^ are listed. n.a., not applicable, n.d., not determined, n.s. no significant difference, *p<0.05.

For each participant, basic laboratory parameters, including blood count and frequencies of immunoglobulin subclasses were determined. The serostatus of the participants concerning selected viral pathogens was determined by IgG antibodies to Hepatitis B (HBV) and C (HCV), (CMV, HSV, and VZV. As shown in Figure 3, nearly all individuals had IgG antibody responses to VZV, while 41% of the young adults and 85% of the elderly were positive for HSV (HSV-1 and/or -2) antibodies. In the group of young adults, the majority was CMV-IgG negative, while the opposite was the case in the elderly (35% vs. 58% positive). 7% of the elderly showed a humoral response to HBV (anti-HBc antibodies), which was absent in young adults, and only a few cases had antibodies to HCV. Since the SARS-CoV2 pandemic occurred during the recruitment of the cohort, and most participants were recruited before the start of the vaccination campaign, only one third had detectable IgG to Spike protein (data not shown), and only 10 donors were convalescent at the time of recruitment. Further on, the participants can be stratified into subgroups regarding influenza, chickenpox, herpes zoster, and human papillomavirus (HPV) vaccinations according to interview responses (Supplemental Figure 1 A, B).

**Figure 3:**
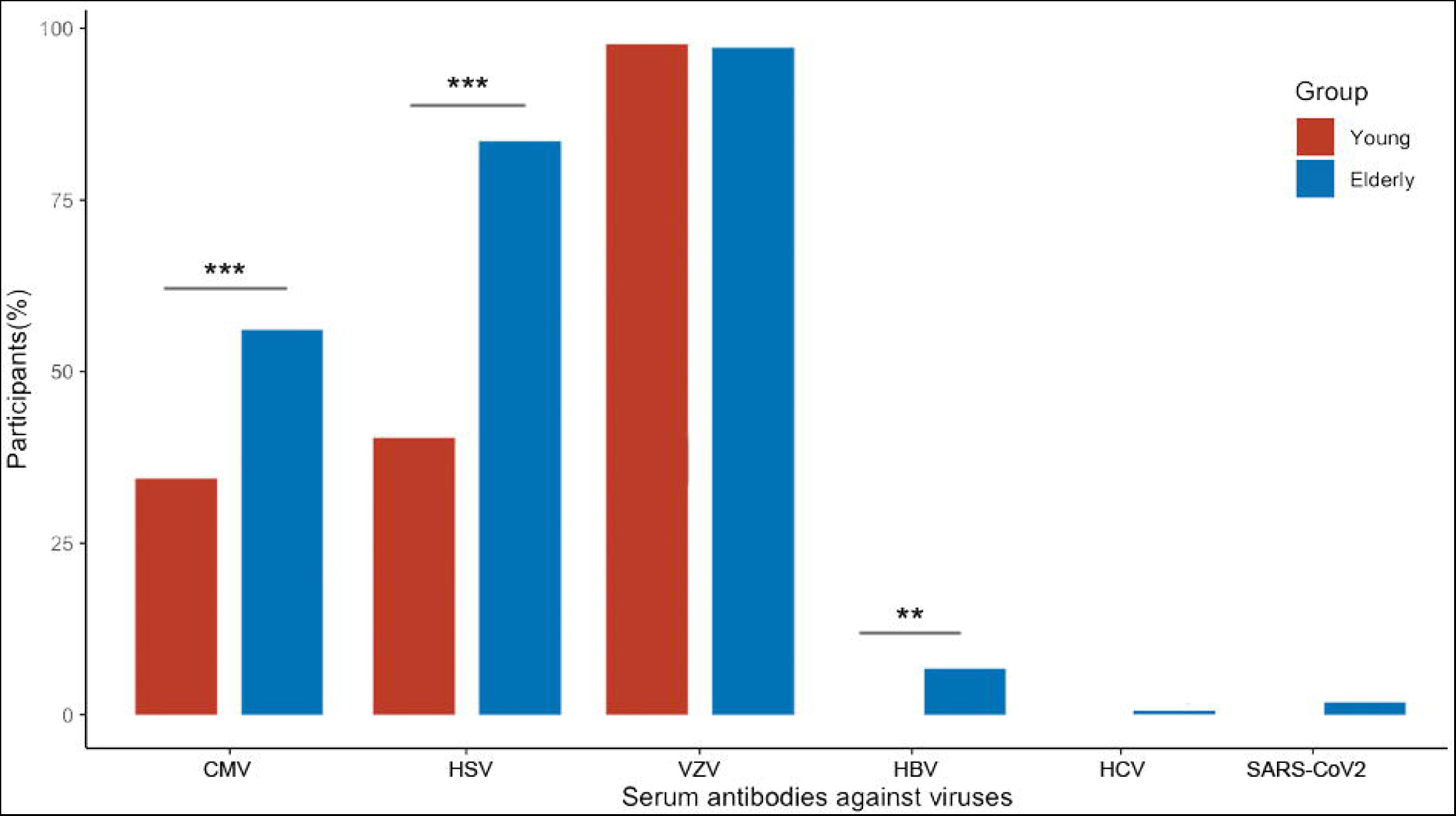
Frequency of participants with serum antibodies against viruses as indicated: CMV: CVM-IgG, HSV: HSV-1/2-IgG, VZV: VZV-IgG, HBV: antibodies against Hepatitis-B-Core-Antigen, HCV: Anti-HCV assay for detection of antibodies against Hepatitis C virus, SARS-CoV2: SARS-CoV2 nucleocapsid protein (NCP)-IgG. Notably, recruitment started before the pandemic, and most participants were recruited before SARS-CoV2 vaccine development. Two-tailed χ² test, **p<0.01, ***p<0.001.

Immediately after blood collection, a blood count was taken and adding to that, the absolute number of T helper cells, cytotoxic T cells, B cells and NK cells was determined by flow cytometry. Absolute blood counts showed a similar pattern as in former reports on immune aging, such as reduced numbers of CD45^+^ leukocytes, total T cells, CD4^+^ T cells, CD8^+^ T cells, and B cells (Figure 4) in elderly compared to young adult donors. Numbers of NK cells have been previously reported to be elevated in older individuals^32^, and we observed a similar trend in our data. High-dimensional flow cytometry with cryopreserved PBMC was carried out in a second step (not shown).

**Figure 4:**
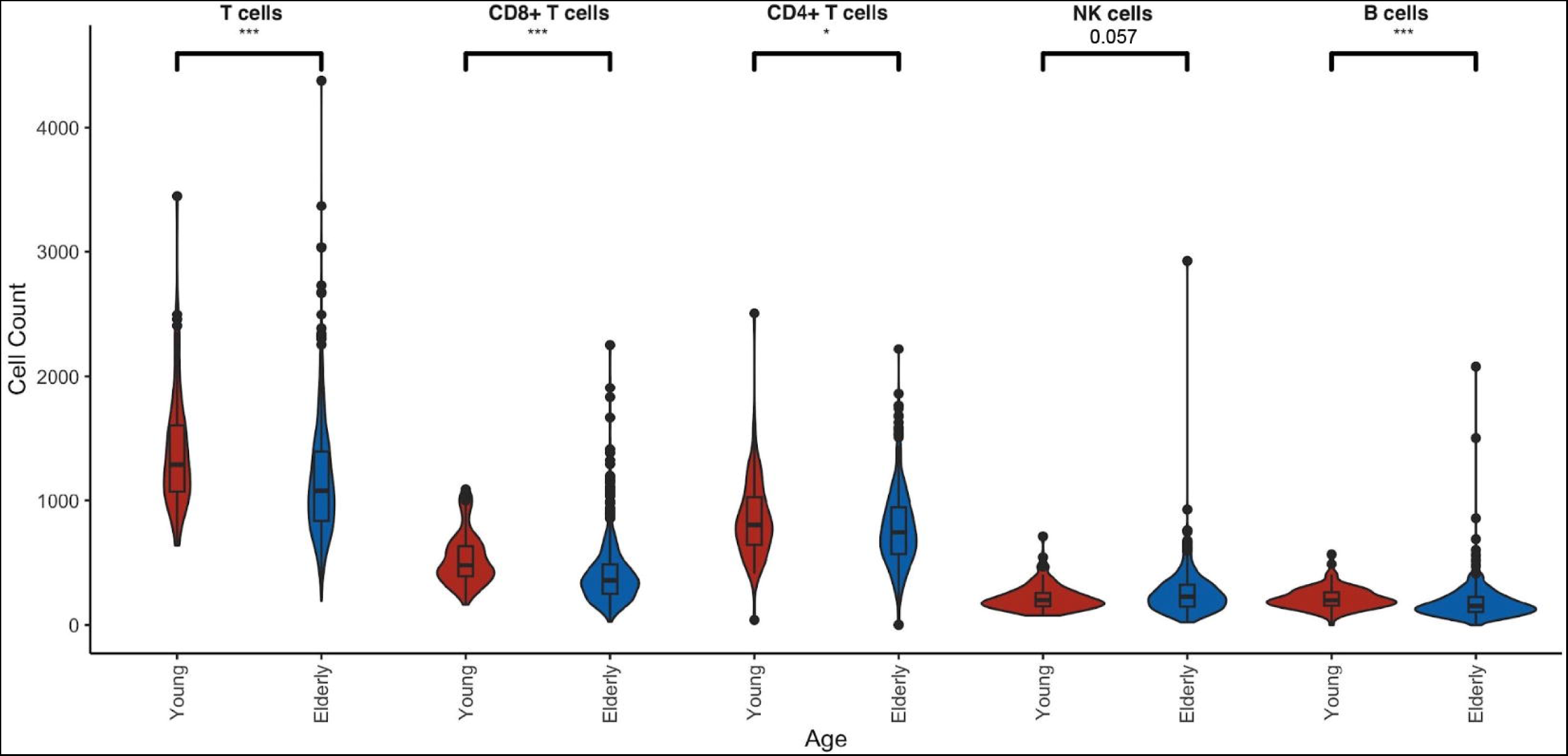
Absolute counts of leukocytes (CD45^+^), T cells (CD45^+^CD3^+^), CD4^+^ T cells (CD45^+^CD3^+^CD4^+^CD8^-^), CD8^+^ T cells (CD45^+^CD3^+^CD4^-^CD8^+^), B cells (CD45^+^CD3^-^CD19^+^), and NK cells (CD45^+^CD3^-^CD16^+^CD56^+^) per µl blood measured by Trucount flow cytometry directly after blood collection. Young adults (20-39 years of age n=99) are compared to the elderly (>60 years of age, n=549). Two-tailed Wilcoxon signed rank test, *p<0.05, **p<0.01, ***p<0.001.

### Next steps and Outlook

To gain a comprehensive picture of how immune responses vary amongst individuals, the biomaterials of each participant are currently analyzed by a multi-omics approach (compare ^33,34^), which includes the layers of genomics, transcriptomics, epigenomics, metagenomics, and metabolomics. These data are extended by cellular phenotyping by high-dimensional flow cytometry and the innate and adaptive immune responses to aforementioned 13 different viral, bacterial and fungal pathogen-derived stimuli. Data of the different -omics layers will be integrated where appropriate to gain detailed insights. This in turn will help us to unmask how host genetic and non-genetic factors influence immune responsiveness, refine our understanding of immune system variation and understand the source of the variability in disease susceptibility between individuals.

### Connected disease cohorts

In parallel to the recruitment of the SI cohort, two patient cohorts were built with a focus on infections by latent virus-reactivation.

In the RESIST VZV cohort, 200 hospitalized patients in the acute disease stage have so far been recruited from December 2019 to March 2024. A telephone interview was performed after six weeks and a follow-up visit took place after 3 months. VZV reactivation is primarily observed with increased age, one of the most frequent infectious diseases in dermatology and neurology, and here the second most frequent cause for encephalitis or meningitis^7^. Although individuals of every age can be affected, elderly individuals are at highest risk^35^.

The RESIST HSV/AD cohort recruits patients suffering from severe, disseminated or systemic HSV-1 infections predominantly occurring in patients suffering from atopic dermatitis (AD), termed eczema herpeticum (EH)^36^. If not treated immediately and properly, these can lead to life-threatening encephalitis, meningitis, and disseminated infections. Current studies suggest that the chronic immune stimulation in these patients contributes to the increased susceptibility to EH^37,38^ and other viral diseases (e.g. eczema coxsackium, eczema vaccinatum, severe chicken pox/herpes zoster). Recruitment started in December 2019 and is still ongoing. Of note, patients in the acute disease stage and also patients with a history of EH are included, as well as patients with AD, who never experienced EH. Patients who are included in the study with active disease can participate in a follow-up visit after 3 months.

Laboratory and clinical findings and also data from the questionnaires from these two patient cohorts will be directly compared to data from the SI cohort. The first 150 recruited patients in each patient cohort mirror the SI cohort with regard to the genome-wide analysis of genetic polymorphisms, and the innate and adaptive immune responses of freshly isolated PBMC to 13 pathogen-derived stimuli (including VZV and HSV-1), which are examined using multi-omics approaches. Moreover, antigen-specific T cell responses to the respective pathogens are measured.

Together, the data of the SI cohort and the VZV and HSV/AD cohorts are used to monitor the immune response and the susceptibility to virus infection/reactivation by measuring and characterizing the individual immune response and cell composition.

## Discussion

We recently recruited a cohort of healthy volunteers in Germany with a focus on advanced age. The data shown in the present manuscript reflect an initial evaluation and characterization of the SI cohort. To qualify as a model for the proportion of the population with an older age, the data collected and analyzed should be in agreement with earlier reports in the literature.

The MoCA is commonly used to evaluate multiple cognitive domains in elderly individuals or to screen patients with mild cognitive complaints^39^. The elderly individuals of the SI cohort had an average MoCA score of 25.7 (± 3.3), and 37.9% scored below the cut-off of 26/30 points. This largely reflects a previously mentioned German-speaking group of 283 individuals aged 65 years or older^29^. More specifically, the elderly individuals in the SI cohort had a slightly lower score (unpaired t-test, p=0.075) and a higher frequency below the threshold by trend (χ² p=0.053). TUG measurement, as a measure of physical fitness and risk of falling, yielded comparable values to those reported in a meta-analysis, when the advanced age of our study participants is taken into consideration^30^. Taken together, these comparisons allow us to conclude that the SI cohort is representative of the older population in Central Europe.

The same conclusion can be drawn from the frequency of infectious diseases reported by our study participants, which was higher compared to the GNC. Thereby, diseases characteristic for an advanced age, like Herpes zoster, which was encountered with a mean age of 55 in the elderly SI cohort participants, need to be investigated separately from those, which appear as a function of longer lifetime. As herpes zoster was reported in the GNC at rates similar to the younger adults in the SI cohort, the elderly of the SI cohort (≥ 60 years) developed the disease at 2-fold higher frequencies. In any case, the SI cohort has the potential to investigate the impact of such a history of infectious disease and the immunological scar it may leave.

The composition of immune cells changes with age, partly as a result of thymus involution and repeated exposure to antigens and pathogens^40^. Repertoire shrinkage and absolute reduction of T cells is a hallmark of the aged immune system, as is the frequency of NK cells, which have been reported to increase in parallel, albeit with reduced cellular mobility and activity^32^. The data of the SI cohort analyzed so far mirror these observations, and future studies using high-dimensional flow cytometry will allow deeper insights into respective immune cell subsets with regard to infection risks in elderly individuals.

Serology testing for HSV and CMV confirmed that the majority of the population gets infected during lifetime. The nature of herpesviruses, with latent infection and constant triggering of the immune system after (unsuccessful) reactivation, is known to have a significant impact on the immune system.

The comparatively high frequency of influenza vaccination among the elderly and, even more convincingly, the rate of multiple vaccinations reflects recent and current vaccination campaigns in Germany. However, Chickenpox vaccination coverage is surprisingly low in the SI cohort, as the Oka strain vaccine was already developed in the 1970ies^41^, but this may be explained by the fact that childhood vaccination was not recommended in Germany until 2004. On the other hand, herpes zoster vaccination with Shingrix is recommended from the age of 60 years onwards, but the vaccine appears to be not well accepted by the population^42^, as mirrored by the SI cohort. This suggests that VZV will remain a common cause of morbidity among elderly individuals. A history of HPV vaccination was found only in the younger participants, in line with the more recent approval of the vaccine in 2006 and the recommendation for its use in 2007.

The in-depth characterization of the subjects and the breadth of the preserved biomaterials form the unique feature of this cohort study and allow the identification of risk factors of ageing with the help of multi-omics analyses. Initial insights into the data collected show that the SI cohort reflects known characteristics of the ageing immune system and the associated susceptibility to infection. The SI cohort thus forms a suitable platform for deciphering molecular mechanisms for effective immunity in older people and to identify risk factors in order to better protect this vulnerable population group in the future.

## Data Availability

A committee of RESIST principal investigators forms a supervisory board overseeing general governance questions and project decisions with possible financial impact (the RESIST SI cohort steering committee). With regard to the rights of access to and use of study data and bio-specimens detailed rules have been defined. Scientific research groups are eligible to apply for data and biomaterial by submitting a proposal to the RESIST SI cohort steering committee. Further information can be received upon contacting the RESIST SI cohort via the BBMRI-ERIC Directory.

## Acknowledgements

This project was funded by the Deutsche Forschungsgemeinschaft (DFG, German Research Foundation) under Germany’s Excellence Strategy – EXC 2155 “RESIST” – Project ID 390874280.

We wish to thank all the participants and patients who volunteered for this study. This study would not have been possible without the work of a large group of staff members from the different departments. In particular, we acknowledge the contributions of Ella Fiebig, Petra Kienlin and Diana Kleppe.

## Author information

### Consortium

The RESIST SI Cohort Investigators consist of the persons named in the authors list and the following persons: Berislav Bošnjak and Rodrigo Gutierrez Jauregui, Institute of Immunology, Hannover Medical School (MHH), Hannover, Germany; Felix Jenniches, Department for Epidemiology, Helmholtz Centre for Infection Research (HZI), Braunschweig, Germany; Norman Klopp, Hannover Unified Biobank (HUB), Hannover Medical School (MHH), Hannover, Germany; Till Robin Lesker, Department of Microbial Immune Regulation, Helmholtz Centre for Infection Research (HZI), Braunschweig, Germany; Martin Stangel, Department of Neurology, Hannover Medical School, Hannover (MHH), Germany, and Department of Translational Medicine Neuroscience, Novartis Institute for BioMedical Research, Basel, Switzerland..

### Contributions

Study design: R.F., J.Hu., T.I., Y.K., B.L., Y.L., L.Ro, T.F.S., M.S., T.S., T.W. Recruitment: S.C., Y.K., B.L., S.T. Biobanking and data management: S.C., J.He., M.G., T.I., F.J., X.J., N.K., V.K., N.v.U. Experiments: L.Ro. Data analysis: M.G., L.Ri., L.Ro. Data interpretation: R.F., M.G., J.H., L.Ri., L.Ro., T.F.S., T.W. Writing: S.C., M.G., Y.K., B.L., L.Ro., T.W. Editing and revising: all authors. Supervision: T.W.

### Additional information

Supplementary information is available for this paper.

Correspondence and requests for materials should be addressed to Thomas Werfel or Lennart Roesner.

## Online Methods

### Governance and data availability

A committee of RESIST principal investigators forms a supervisory board overseeing general governance questions and project decisions with possible financial impact (the RESIST SI cohort steering committee). With regard to the rights of access to and use of study data and bio-specimens detailed rules have been defined. Scientific research groups are eligible to apply for data and biomaterial by submitting a proposal to the RESIST SI cohort steering committee. Further information can be received upon contacting the RESIST SI cohort via the <u>BBMRI-ERIC Directory.</u>

### Ethics and data confidenciality

The SI cohort is being conducted in compliance with all pertinent legislation and directives and following the guidelines on human biobanks for research and other relevant directives on research ethics. Special emphasis is placed on privacy, safety of data, genetic information and reporting of incidental medical findings to participants. Ethics committee of Hannover Medical School (MHH), Hannover, Germany, gave ethical approval for this work (No 8615_BO_S_2019; connected disease cohorts under the file references 8730_BO_S_2019 and 8733_BO_S_2019). The study was conducted according to the principles of declaration of Helsinki. Participants gave their written informed consent prior to the study.

### Virus serology

Serum samples were tested in the routine diagnostic of the clinical virology lab at MHH. Serology was performed on the Architect System from Abbott Diagnostics (Abbott GmbH & Co. KG, Wiesbaden, Germany) using the Anti-HBc II test for detection of antibodies against the Hepatitis-B-Core-Antigen (HBV), the Anti-HBs test to determine the vaccination titre, the Anti-HCV assay for detection of antibodies against Hepatitis C virus (HCV-AB) and CMV IgG assay for detection of IgG antibodies against CMV. The Detection of IgG antibodies for VZV and HSV was done on the Liaison XL analyzer from Diasorin (DiaSorin S.p.A. Saluggia (VC), Italy) using the LIAISON® VZV IgG and the LIAISON® HSV-1/2 IgG assay. The detection of IgG antibodies against SARS-CoV-2 nucleocapsid protein (NCP) and Spike were done on the EUROIMMUN Analyzer I using the Anti-SARS-CoV-2-NCP-ELISA (IgG) and the Anti-SARS-CoV-2-ELISA (IgG), respectively (Euroimmun, Lübeck, Germany).

### Absolute cell count and basic immunophenotyping

Whole fresh blood was subjected to FACS Lysing Solution (#349202, BD Biosciences, Franklin Lakes, NJ, USA) and subsequently the 6-color TBNK Reagent with TruCount tubes (#337166, BD Biosciences) according to the manufacturer’s protocol (Supplemental Figure 2). Measurements were taken from distinct samples. Fluorescence signals and TruCount beads were acquired on a 3-laser CytoFLEX using Cytexpert software 2.3 (Beckman Coulter, Brea, CA, USA). Instrument quality control and standardization were performed daily using CytoFLEX Daily QC Fluorospheres (Beckman Coulter #B53230).

### Statistics

Group comparisons between two groups were conducted using the Wilcoxon signed rank test as indicated. Associations between two-factors were assessed by x² test as indicated in each figurés or tablés legend. P values ≤0.05 were considered statistically significant.

Analyses were conducted with GraphPad Prism (version 5.02, GraphPad, Boston, MA, USA) or R (version 4.3.2).Tables

