## Supplemental Figures for "The RESIST Senior Individuals Cohort: Design, participant characteristics and aims"

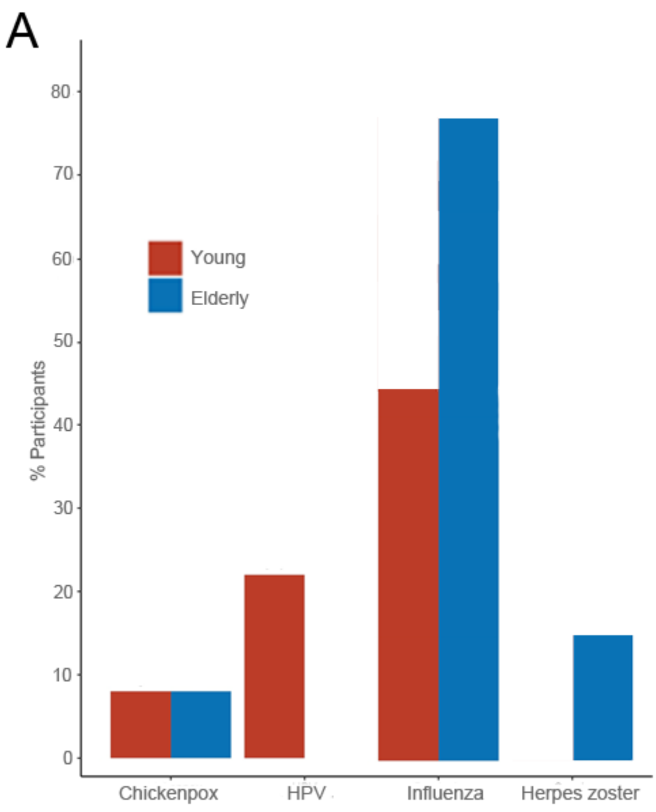


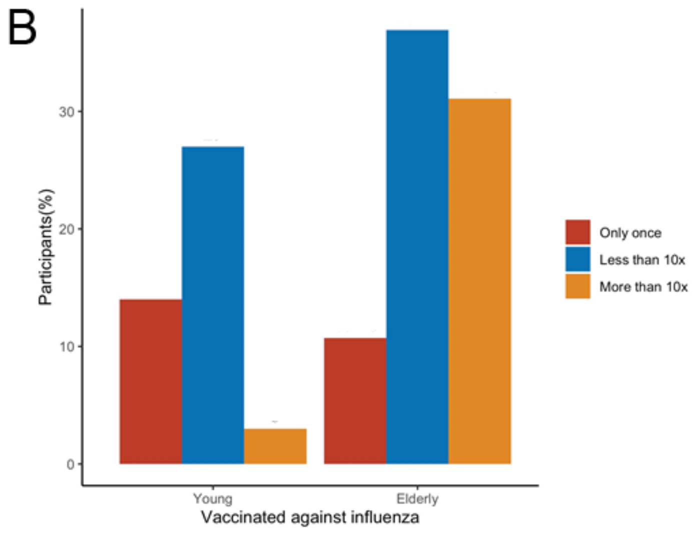


**Suppl. Figure 1**: (A) Frequency of participant-reported vaccination(s) against Chickenpox, Human papilloma virus, Influenza, and Herpes zoster. (B) Participant-reported vaccination(s) against Influenza in detail.

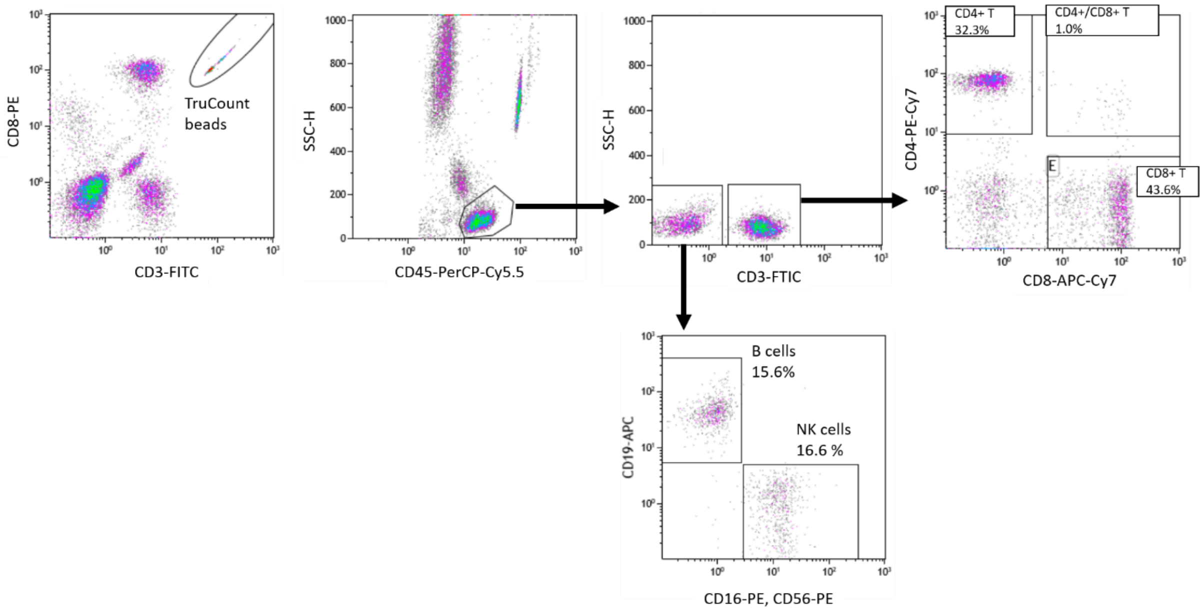


**Suppl. Figure 2**: Gating strategy of the basic immunophenotyping.
